# Utility of quantitative measurement of *T_2_* using Restriction Spectrum Imaging for detection of clinically significant prostate cancer

**DOI:** 10.1101/2024.03.29.24305033

**Authors:** Mariluz Rojo Domingo, Christopher C Conlin, Roshan A Karunamuni, Courtney Ollison, Madison T Baxter, Karoline Kallis, Deondre D Do, Yuze Song, Joshua M Kuperman, Ahmed S Shabaik, Michael E Hahn, Paul M Murphy, Rebecca Rakow-Penner, Anders M Dale, Tyler M Seibert

## Abstract

**Background:** The Restriction Spectrum Imaging restriction score (RSIrs) has demonstrated higher diagnostic accuracy for clinically significant prostate cancer (csPCa) than conventional DWI. Both diffusion and *T_2_* properties of prostate tissue inform the RSI signal, and studies have shown that each may be valuable for csPCa discrimination.

**Purpose:** To determine whether prostate *T_2_* varies across RSI compartments and in the presence of csPCa, and to evaluate whether consideration of compartmental *T_2_* (c*T_2_*) improves csPCa detection over RSIrs alone.

**Study Type:** Retrospective.

**Population:** Two cohorts (46 and 195 patients) scanned for csPCa.

**Field Strength/Sequence:** 3T multi-*b*-value DWI acquired at multiple TEs.

**Assessment:** c*T_2_* values were computed from multi-TE RSI data and compared between RSI model compartments. csPCa detection was compared between RSIrs and a logistic regression model (LRM) for predicting the probability of csPCa using c*T_2_* in combination with RSI measurements.

**Statistical Tests:** *T*wo-sample t-tests (α=0.05) were used to compare c*T_2_* values between compartments and between patients with and without csPCa. Area under the receiver operating characteristic curve (AUC) was used to evaluate csPCa detection performance.

**Results:** In both cohorts, *T_2_* differed (*p*<0.05) across all RSI compartments (*C_1,_ C_2,_ C_3,_ C_4_*). Voxel-level data from cohort 1 showed that *T_2_* differed between normal and cancerous tissue in *C_1,_ C_2,_ C_3_* (*p*<0.001). Whole-prostate *T_2_* differed between patients with and without csPCa in *C_3_* (*p*=0.02). In cohort 2, whole-prostate *T_2_* differed in *C_1_* (*p*=0.01), *C_3_* (*p*=0.01), and *C_4_* (*p*<0.01). Consideration of c*T_2_* improved csPCa discrimination compared to diffusion alone, but not compared to RSIrs [cohort 1: 0.80 vs 0.70 (diffusion) and 0.80 (RSIrs), cohort 2: 0.72 vs 0.65 (diffusion) and 0.72 (RSIrs)].

**Data Conclusion:** Significant differences in c*T_2_* were observed between normal and cancerous prostatic tissue. With our data, however, consideration of c*T_2_* did not significantly improve cancer detection performance over RSIrs alone.

## Introduction

Multiparametric magnetic resonance imaging (mpMRI) has become an important tool for the diagnosis of prostate cancer (PCa)^1^. MpMRI has proven to reduce unnecessary biopsies, mitigates overdiagnosis of clinically insignificant prostate cancer (indolent PCa), and enhances detection of clinically significant prostate cancer (csPCa, Grade Group 2 or higher)^2,3^. In standard reporting of prostate MRI (Prostate Imaging Reporting and Data System, PI-RADS v2.1^4^), diffusion-weighted imaging (DWI) and *T_2_* -weighted imaging are the principal modalities used to detect csPCa. DWI measures the random movement of water molecules within tissues, aiding in the visualization of areas of restricted diffusion associated with hypercellular csPCa^5^. Meanwhile, *T_2_* -weighted imaging provides detailed anatomical information and facilitating the visualization of abnormalities in prostate tissue. The combined analysis of these two sequences allows for a more in-depth evaluation of potential tumor lesions, thereby contributing to csPCa detection and characterization.

The challenge with the interpretation of conventional mpMRI lies in its inherent subjectivity and variability^6^. The interpretations of imaging data by different radiologists that rely on qualitative assessment alone leads to inconsistencies in the identification and characterization of csPCa lesions^7^. Interobserver variability significantly limits the accuracy and reliability of csPCa diagnosis^8^. To enhance diagnostic accuracy, there is a growing emphasis on the development and adoption of quantitative MRI approaches.

Quantitative MRI aims to provide objective metrics of tissue properties associated with the probability of csPCa, offering the potential for more standardized and reproducible image assessment.

Restriction Spectrum Imaging (RSI) is a quantitative approach to DWI for csPCa detection and characterization. RSI scans are acquired at multiple *b*-values (diffusion weightings) to distinguish diffusion signal from tissue micro-compartments (intracellular water, extracellular hindered water, freely diffusing water, and flowing fluid)^9–12^. However, RSI models typically do not incorporate quantitative *T_2_* measurements. On the other hand, studies using luminal water imaging (LWI) and hybrid multidimensional MRI have shown that tissue *T_2_* can differ between prostate tissue compartments and provide diagnostic information that is complementary to diffusion^13,14^.

In this study, we acquired prostate RSI data at multiple echo times (TEs) to measure compartmental *T_2_* in addition to diffusion. We aimed to determine whether compartmental *T_2_* (i.e., *T_2_* within each RSI micro-compartment) differs between cancerous and normal prostate tissue, and whether consideration of compartmental *T_2_* in RSI yields improved detection of csPCa.

## Methods

### Study Population

This study was approved by the institutional review board (IRB). For the prospective cohort, a waiver of consent was obtained to acquire images with two TEs for image optimization, as well as to access patient images and other clinical information. Consecutive patients of age 18 years or older who underwent MRI with multi-TE RSI for suspected PCa were included in the two cohorts of this study. The first cohort (retrospective) included patients scanned for PCa between August and December of 2016 with multi-TE RSI as part of a quality improvement project. The second cohort included patients who were scanned for PCa between March 2021 and January of 2023. Additionally, the second cohort included participants in a separate, prospective study (also approved by the IRB) of patients with localized prostate cancer using the same multi-TE RSI protocol. These latter participants gave informed consent to undergo additional imaging before and after radiation therapy; only pre-treatment imaging was included in the present study. Patients were excluded if they had received any treatment for PCa prior to the MRI acquisition or if a lesion with PI-RADS score ≥3 was detected on MRI but no biopsy information was available.

### Routine Clinical Evaluation

Most patients in both cohorts underwent prostate MRI as part of routine clinical care for PCa. MpMRI was performed according to PI-RADS guidelines, and interpretation was made per clinical routine using PI-RADS v2.1. Several patients, all from cohort 2, had PCa diagnosed on systematic biopsy without MRI, and then had an MRI with RSI before any treatment as part of a prospective study; PI-RADS scores are not available for these subjects. The presence of clinically significant prostate cancer (csPCa, grade group 2 or higher) was determined from the best available histopathology, either biopsy (typically systematic 12-core biopsy with additional targeted cores for suspicious lesions on MRI) or radical prostatectomy. Patients with PI-RADS lesions of 1 or 2 with no biopsy were considered negative for csPCa, in accordance with European Association of Urology (EAU) guidelines^15–17^.

### RSI Data Acquisition and Processing

All patients were scanned with an expanded MRI protocol that included two multi-*b*-value RSI acquisitions performed with different TEs. MRI acquisition details are summarized in Table 1. All MR imaging was performed on a 3T clinical scanner (Discovery MR750; GE Healthcare, Waukesha, WI, USA), using a 32-channel phased-array coil over the pelvis. For each patient, two axial, multi-*b*-value DWI volumes were separately acquired using two different TEs: 80 ms and 100 ms for cohort 1, and 76 ms and 90 ms for cohort 2. The other parameters were the same between scans. In addition to the DWI volumes, a single *T_2_*-weighted volume was acquired for anatomical reference using the same scan coverage as the DWI volumes. MRI post-processing was performed using programs implemented in MATLAB (MathWorks, Natick, MA, USA). DWI volumes were corrected to account for *B_0_*-inhomogeneities, gradient nonlinearities, eddy currents^18^, and image noise^10^. Samples at each *b*-value were averaged together. Image registration^19^ was applied to correct for patient motion between acquisitions.

**Table 1:** Acquisition details for DWI and *T_2_*-weighted image volumes. All MR imaging was performed on a 3T clinical scanner (Discovery MR750; GE Healthcare), using a 32-channel phased-array coil over the pelvis. The two DWI volumes for both cohorts were acquired using different TEs to allow for examination of *T_2_* relaxation in prostatic tissue compartments. The single *T_2_*-weighted volume was acquired for anatomical reference.

| Cohort 1 | DWI 1 | DWI 2 | $T_2$-weighted |
| --- | --- | --- | --- |
| Pulse sequence | Diffusion-weighted EPI* | Diffusion-weighted EPI* | FSE† |
| TR (ms) | 5000 | 5000 | 6225 |
| TE (ms) | 80 | 100 | 100 |
| FOV (mm) | 220 | 220 | 220 |
| Matrix<br>[resampled dimensions] | 96 x 96 x [128 x 128] | 96 x 96 x [128 x 128] | 320 x 320 x [512 x 512] |
| Slices | 34 | 34 | 34 |
| Slice Thickness (mm) | 3 | 3 | 3 |
| $b$-values (s/mm<sup>2</sup>)<br>[number of samples] | 0[7‡], 200 [6], 1000 [6],<br>2000 [6], 3000 [6] | 0[7‡], 200 [6], 1000 [6],<br>2000 [6], 3000 [6] | N/A |

\*Echo-planar imaging
†Fast spin echo
‡An extra $b = 0$ s/mm<sup>2</sup> volume was acquired with reverse phase encoding to enable correction of B0-inhomogeneity distortions

Table 1: Acquisition details for DWI and $T_2$ -weighted image volumes.
| Cohort 2 | DWI 1 | DWI 2 | $T_2$-weighted |
| --- | --- | --- | --- |
| Pulse sequence | Diffusion-weighted EPI* | Diffusion-weighted EPI* | FSE† |
| TR (ms) | 4500 | 4500 | 6230 |
| TE (ms) | 76 | 90 | 98 |
| FOV (mm) | 200 x 100 | 200 x 100 | 200 x 200 |
| Matrix<br>[resampled dimensions] | 80 x 48 x [128 x 128] | 80 x 48 x [128 x 128] | 320 x 320 x [512 x 512] |
| Slices | 32 | 32 | 32 |
| Slice Thickness (mm) | 3 | 3 | 3 |
| $b$-values (s/mm<sup>2</sup>)<br>[number of samples] | 0 [2‡], 50[6], 800[6],<br>1500[12], 3000 [18] | 0 [2‡], 50[6], 800[6],<br>1500[12], 3000 [18] | N/A |

\*Echo-planar imaging
†Fast spin echo
‡An extra $b = 0$ s/mm<sup>2</sup> volume was acquired with reverse phase encoding to enable correction of B0-inhomogeneity distortions

For patients in cohort 1, regions of interest (ROIs) were manually defined on *T_2_*-weighted images over the whole prostate, peripheral zone, and transition zone (the central zone was included with the transition zone). The contouring of the prostate zones and tumor lesions was performed using MIM software (MIM Software, Inc; Cleveland, OH, USA), by a radiation oncologist with 3 years of experience and two board-certified sub-specialist radiologists with 4 and 6 years of experience, using all available clinical imaging and pathologic information^11^. Radiologist-certified contours of the prostate zones and lesions were not obtainable for cohort 2. Instead, contours of the whole prostate were generated automatically using OnQ Prostate software from Cortechs Labs (Cortech.ai; San Diego, CA, USA).

### RSI modeling

Prior studies established and validated a four-compartment RSI model of the diffusion _signal10,11,20:_

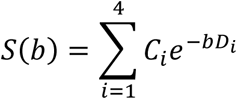

*S(b)* denotes the measured DWI signal intensity at a particular *b*-value, which is modeled as a linear combination of exponential decays representing four diffusion compartments. *C_i_* describes the compartmental signal contributions to be determined via model-fitting. The diffusion coefficients, *D_i_*, are fixed for each of the four tissue compartments to empirically determined^10^ values that broadly represent restricted, hindered, free diffusion, and vascular flow: 1.1e-4, 1.8e-3, 3.6e-3, and 0.1220 mm^2^/s, respectively. Signal-contribution (*C_i_*) maps were computed for both DWI volumes per patient by fitting this model to the signal-vs.-*b*-value curve from each voxel. A previously validated biomarker for PCa called the RSI restriction score (RSIrs) was computed by dividing the signal intensity of the restricted diffusion compartment, *C_1_*, at each voxel by the median signal intensity within the whole prostate on the *b* = 0 mm^2^/s DWI images (an index of apparent *T_2_*-weighting in the prostate)^11,20–23^.

### Compartmental *T_2_* mapping and analysis by csPCa status

*T_2_* maps were computed for each compartment of the RSI model by fitting the monoexponential *T_2_* decay formula to the signal values from the two *C_i_* maps with different TEs. The median *T_2_* within the whole prostate was then computed. Two-sample *t*-tests (*α* = 0.05) were used to determine whether there were significant differences in median *T_2_* between compartments.

For cohort 1, two-sample *t*-tests (*α* = 0.05) were used to compare median compartmental *T_2_* values between benign or clinically insignificant PCa tissue and csPCa lesions. For both cohorts, we used two-sample *t*-tests (*α* = 0.05) to compare each patient’s median *T_2_* by compartment in the whole prostate, and whether compartmental *T_2_* was significantly different between patients with and without csPCa. Any compartments with a significant difference in *T_2_* between normal and cancerous tissue were noted for inclusion in subsequent multivariable modeling.

### Logistic regression model fitting and evaluation of cancer-detection performance

A logistic regression model (LRM) was developed to estimate the probability that a given voxel of tissue contains csPCa given measurements of diffusion and compartmental *T_2_*. RSIrs^11^ was included as the diffusion parameter of the model. Compartmental *T_2_* was included in the LRM for each compartment that showed a significant difference in *T_2_* between normal and cancerous tissue. Cohort 1 had radiologist-certified lesion contours available and was therefore used to train the LRM. In patients with csPCa, voxels inside the lesion contours were labeled as csPCa-positive, while prostate voxels outside the lesion contours were labeled as csPCa-negative. In these patients, voxels labeled as csPCa-positive were included to train the LRM and all non-csPCa voxels were excluded. In patients without csPCa, diffusion and compartmental *T_2_* measurements from all voxels within the entire prostate were used to train the LRM and labeled as csPCa-negative.

Ten-fold cross-validation was performed to evaluate voxel-level csPCa-detection performance of the model within cohort 1. We assessed csPCa-detection performance using the area under the receiver operating characteristic curve (AUC) and calculated 95% confidence intervals (CI) from 10,000 bootstrap samples.

Both cohorts were used to test the patient-level csPCa-detection performance of the model. For patient-level analysis of the LRM, the highest probability value observed within the whole prostate was used as the predictor variable. Similarly, the maximum RSIrs value within the whole prostate was used as the patient-level predictor for RSIrs. We also computed maximum *C_1_* to obtain the patient-level performance of diffusion only, as RSIrs incorporates global prostate *T_2_* signal in addition to diffusion signal. AUC values were computed for maximum *C_1_*, maximum RSIrs, and the LRM, and compared using two-sample *t*-tests (*α* = 0.05). The 95% confidence intervals were estimated through random sampling with replacement from 10,000 bootstrap patient samples.

## Results

### Study Population

Cohort 1 comprised 46 patients (age: 70 ± 10 years; PSA: 10.6 ±16.9 ng/mL). Cohort 2 comprised 195 patients (age: 69 ± 8 years; PSA: 8.2 ± 8.5 ng/mL). In cohort 1, 22 of 46 patients (47.8%) had csPCa, while the remaining 24 had either low-grade (grade group 1) disease or no cancer. In cohort 2, 96 of 195 (49.2%) patients had csPCa. 38 participants from cohort 2 had no PI-RADS scores available because csPCa was diagnosed on systematic biopsy without MRI and then had an MRI with RSI acquisition as part of a separate, prospective study. Table 2 summarizes the patient characteristics of both cohorts.

**Table 2.** Summary of radiologic and pathologic characteristics of the two cohorts of patients included in this study.

| Cohort 1 (n = 46) |  |  |  | Cohort 2 (n = 195) |  |  |  |
| --- | --- | --- | --- | --- | --- | --- | --- |
| Available Pathology | Systematic biopsy only |  | 6 | Available Pathology | Systematic biopsy only |  | 44 |
|  | Targeted Biopsy Only |  | 0 |  | Targeted Biopsy Only |  | 5 |
|  | Systematic and targeted biopsy |  | 20 |  | Systematic and targeted biopsy |  | 60 |
|  | Prostatectomy |  | 15 |  | Prostatectomy |  | 28 |
|  | No biopsy or prostatectomy* |  | 5 |  | No biopsy or prostatectomy* |  | 31 |
| PI-RADS |  | 1 | 9 | PI-RADS |  | 1 | 66 |
|  |  | 2 | 2 |  |  | 2 | 10 |
|  |  | 3 | 10 |  |  | 3 | 18 |
|  |  | 4 | 10 |  |  | 4 | 36 |
|  |  | 5 | 15 |  |  | 5 | 27 |
|  |  | Not available | 0 |  |  | Not available† | 38 |
| Gleason Grade Group | Benign |  | 17 | Gleason Grade Group | Benign |  | 38 |
|  |  | 1 | 2 |  |  | 1 | 30 |
|  |  | 2 | 9 |  |  | 2 | 40 |
|  |  | 3 | 8 |  |  | 3 | 32 |
|  |  | 4 | 1 |  |  | 4 | 7 |
|  |  | 5 | 4 |  |  | 5 | 17 |
| *4 patients had PI-RADS 1 scores and 1 patient had PI-RADS 2 scores. |  |  |  | *27 patients had PI-RADS 1 scores and 4 patient had PI-RADS 2 scores. |  |  |  |
|  |  |  |  | †Part of a prospective research study without clinical evaluation by a radiologist |  |  |  |

### Compartmental *T_2_* mapping

Figure 1 shows compartmental *T_2_* maps for two patients with csPCa, one from each cohort.

**Figure 1:**
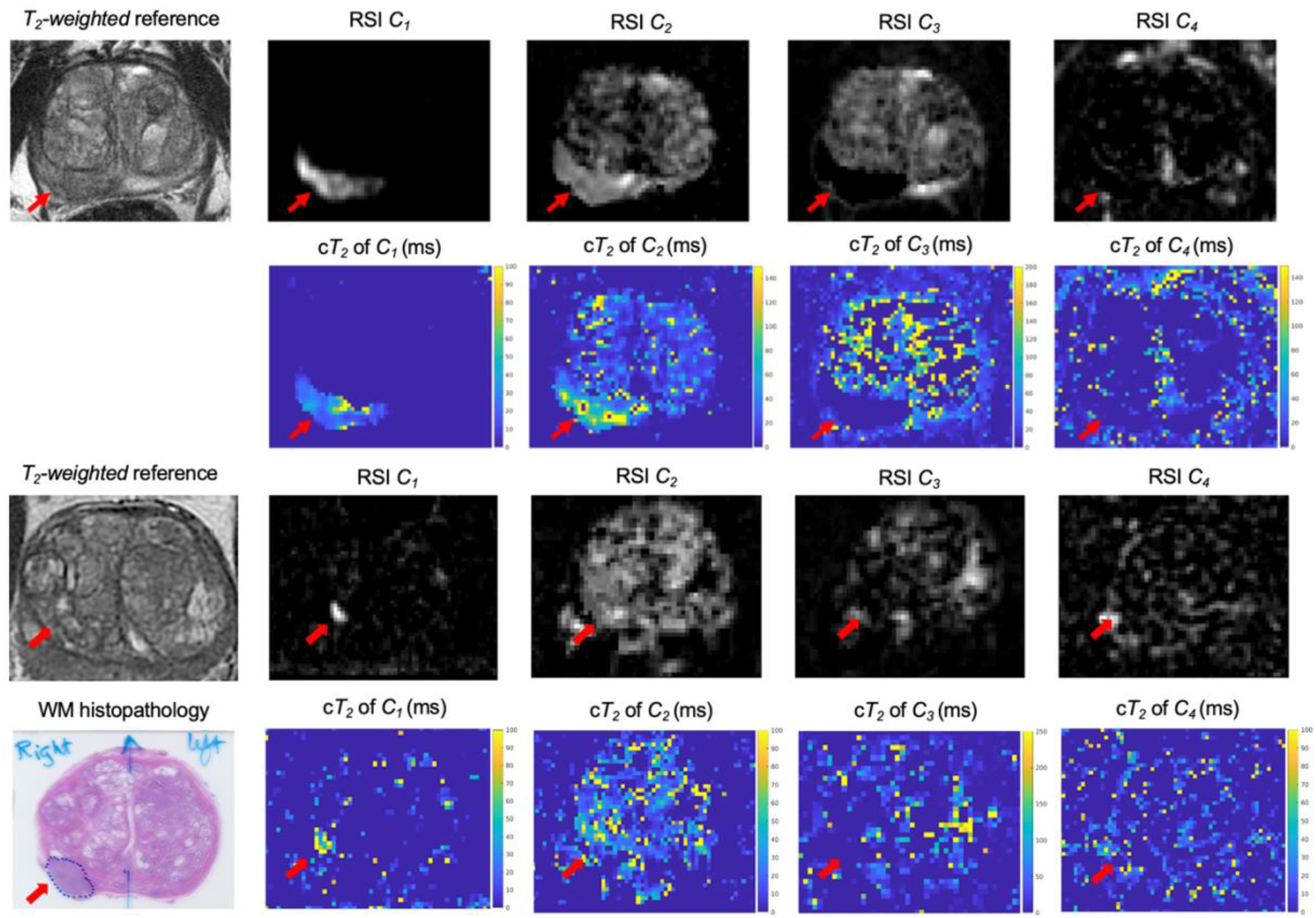
RSI signal contribution (*C_i_*) and compartmental *T_2_* (c*T_2_*) maps for two patients with csPCa. The top panel corresponds to a patient from cohort 1 and the bottom panel to a patient from cohort 2. Compartmental *T_2_* maps were computed for each RSI model compartment by measuring the *T_2_* -weighted signal decay of the signal-contribution map for different TEs. The signal-contribution maps shown here were computed from the DWI acquisition with shorter TE. Whole-mount (WM) histopathology results were available for the patient from cohort 2 and illustrate the lesion contour in the prostate.

Figure 2 shows violin plots of median *T_2_* within the whole prostate for each RSI model compartment. In both cohorts, the highest median *T_2_* values were observed in *C_3_*, followed by *C_2_*, *C_4_*, and finally *C_1_*. Compartmental *T_2_* was significantly different between any two compartments (*p* < 0.05).

**Figure 2:**
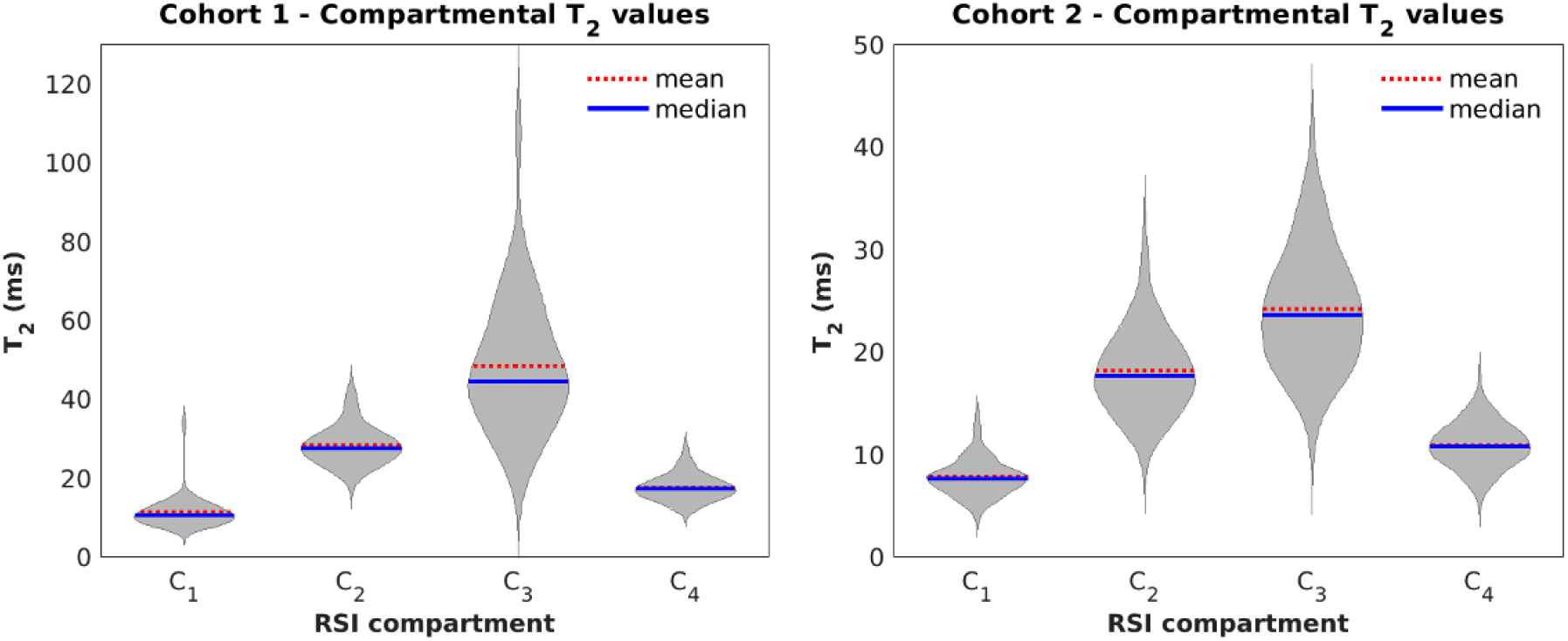
Violin plots showing the distribution of median *T_2_* values in the whole prostate for each of the four RSI model compartments. Within each cohort, compartmental *T_2_* was significantly different between any two compartments (*p* < 0.05). Left panel: cohort 1 (n = 46). Right panel: cohort 2 (n = 195).

### Voxel-level analysis of compartmental *T_2_* values by csPCa status

For each compartment of the RSI model, the comparison of *T_2_* values between csPCa and prostate tissue outside of csPCa lesions is illustrated in Figure 3. This figure corresponds to cohort 1, which has radiologist-certified csPCa lesion contours. csPCa lesions showed significantly higher compartmental *T_2_* values in compartment 1 (*p*-value << 0.001) and compartment 2 (*p*-value << 0.001) than normal tissue. In addition, the compartmental *T_2_* values of compartment *C*_3_ were significantly lower in csPCa lesions (*p*-value << 0.001). The compartmental *T_2_* values for compartment *C*_4_ were not significantly different between csPCa and normal tissues (*p*-value = 0.17).

**Figure 3:**
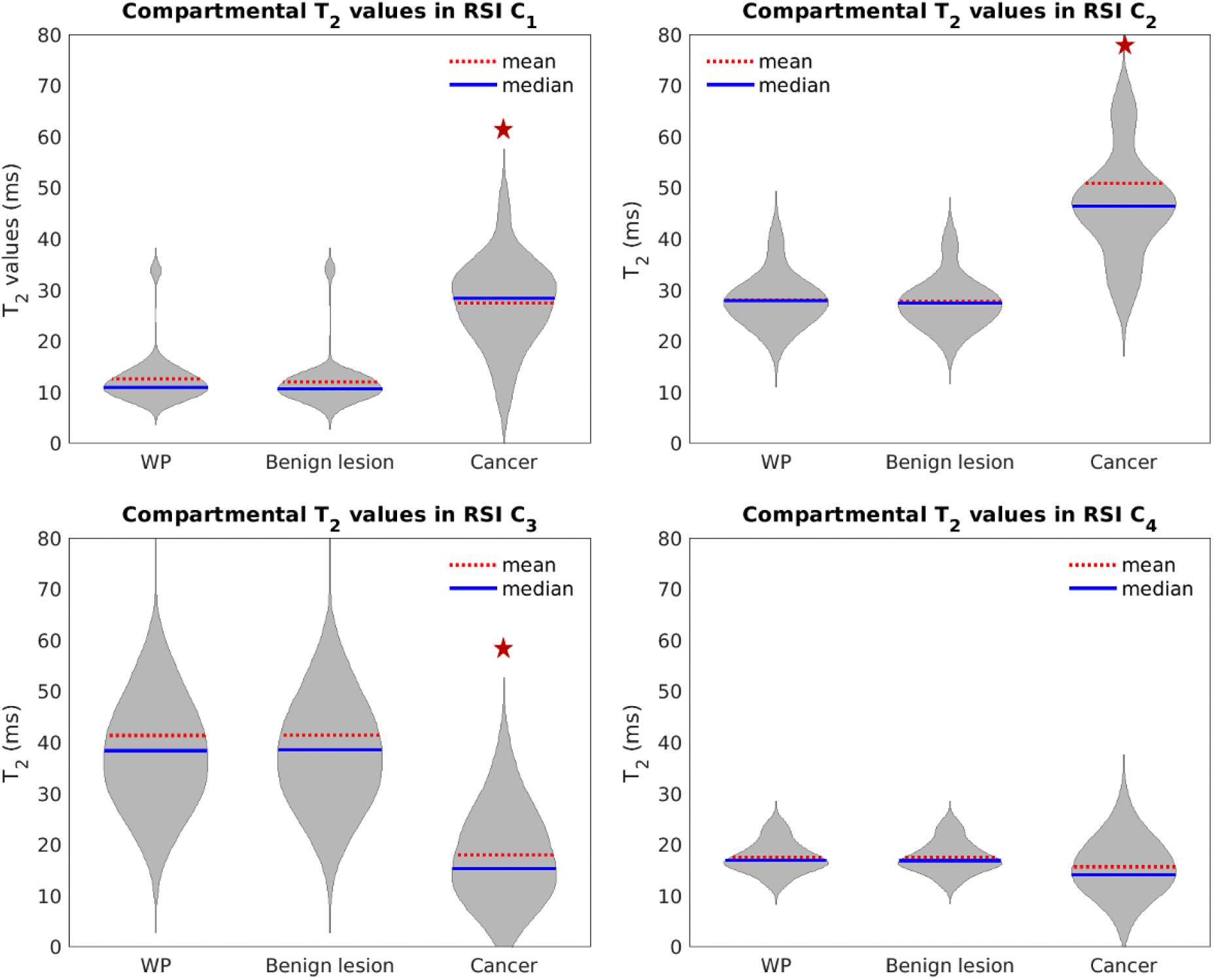
Violin plots showing the distribution of median compartmental *T_2_* values from cohort 1 in both csPCa lesions and the surrounding prostate tissue. The red “+” corresponds to the mean value in each violin. Each panel corresponds to one of the RSI diffusion compartments. csPCa lesions showed significantly higher compartmental T2 values in *C_1_* (*p*-value << 0.001) and *C_2_* (*p*-value << 0.001) than tissues outside of csPCa lesions. Compartmental *T_2_* values of *C_3_* were also significantly lower in csPCa lesions than outside csPCa lesions (*p*-value << 0.001). A red star indicates a significant difference (*p-*value < 0.05) in compartmental *T_2_* between csPCa lesions and the prostate tissue outside the lesions. WP: whole prostate.

### Patient-level analysis of compartmental *T_2_* values by csPCa status

The comparison of compartmental *T_2_* values between patients with and without csPCa is shown in Figure 4. In both cohorts, patients with csPCa had higher compartmental *T_2_* values in compartment *C*_1_ than patients with no csPCa. In cohort 2, median *C_1_* compartmental *T_2_* was significantly higher (*p*-value = 0.07 for cohort 1; *p*-value = 0.01 for cohort 2). Median *C_3_* compartmental *T_2_* was significantly different between csPCa and patients without csPCa in both cohorts (*p*-value = 0.02 for cohort 1; *p*-value = 0.01 for cohort 2). Median *C_4_* compartmental *T_2_* was also significantly different in cohort 2 (*p*-value << 0.01). Compartmental *T_2_* values for the other compartments were not significantly different between patients with and without csPCa.

**Figure 4:**
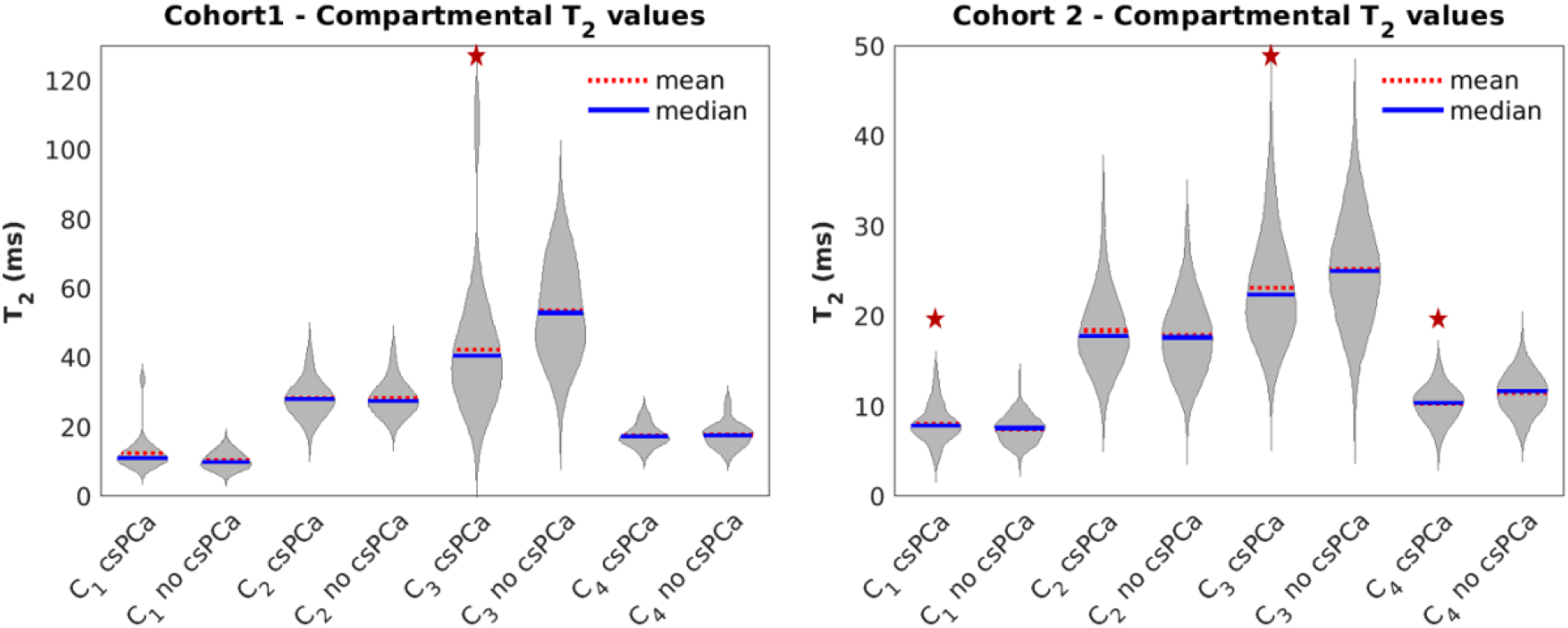
Violin plots comparing compartmental *T_2_* within the whole prostate between patients with csPCa and those without. A red star indicates a significant difference (*p-* value < 0.05) in whole-prostate compartmental *T_2_* between csPCa and non-csPCa patients. Left panel: cohort 1 (n = 46). Right panel: cohort 2 (n = 195).

### Logistic regression model fitting and evaluation of csPCa-detection performance

*T_2_* measurements from RSI compartments 1, 2, and 3 (*C_1_*, *C_2_*, *C_3_*) were included as the *T_2_* parameters of the LRM. These three compartments showed significantly different median *T_2_* signal between in csPCa lesions vs csPCa-negative voxels. The LRM predictors were RSIrs, *C_1_ T_2_* , *C_2_ T_2_*, and *C_3_ T_2_*. The model coefficients with 95% confidence interval for the y-intercept and RSIrs were 6.367 (6.323, 6.416) and -51.621 (-52.354, -50.888), respectively. The weights for *C_1_ T_2_* , *C_2_ T_2_*, and *C_3_ T_2_* were < 0.005. Example probability maps computed from the model are shown in Figure 5 for two patients with csPCa, alongside maps of RSIrs.

**Figure 5:**
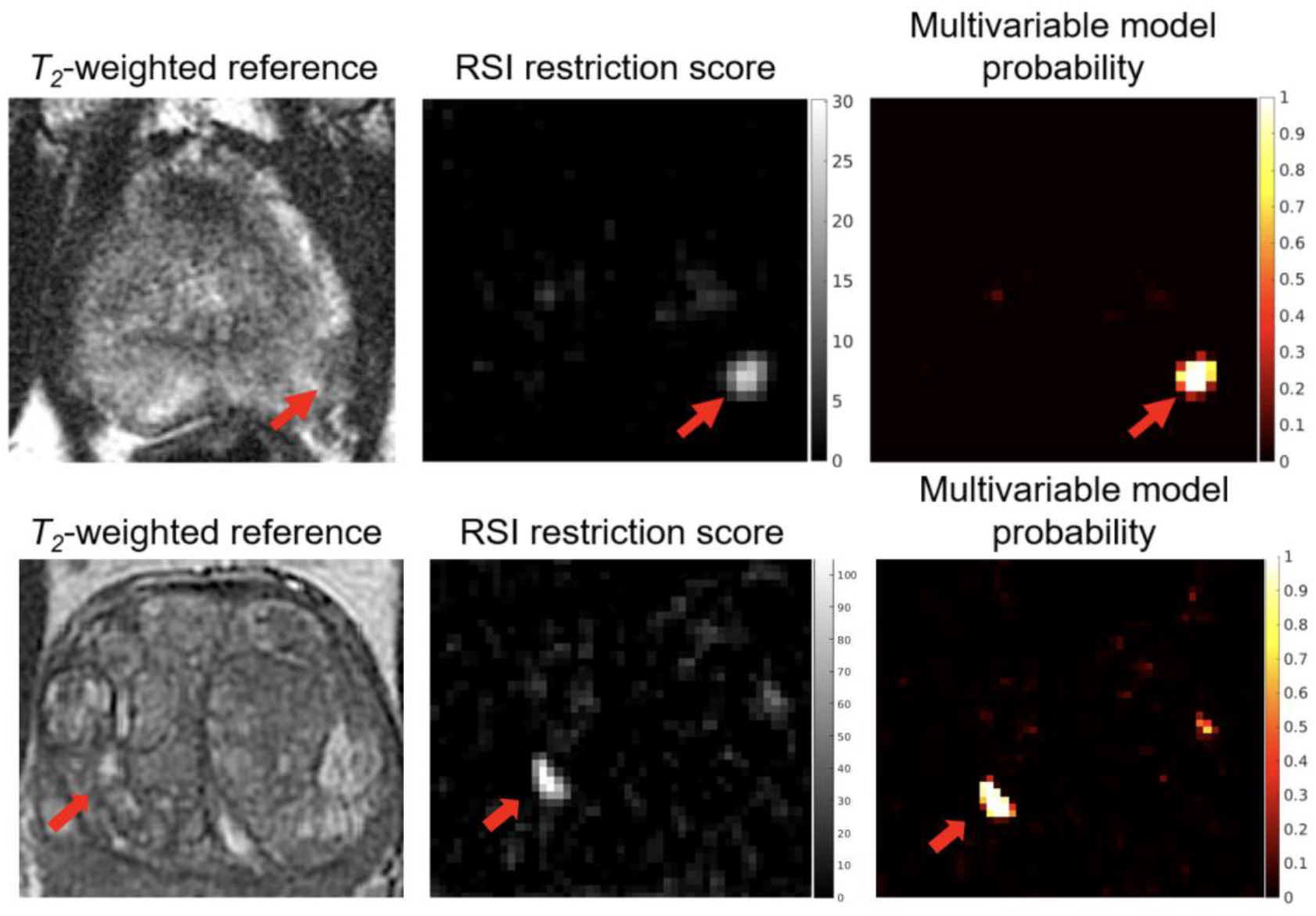
RSI restriction score and multivariable model probability maps of the prostate for patients with csPCa. Top panel: Patient from cohort 1 with a lesion in the left peripheral zone. Bottom panel: Patient from cohort 2 with a lesion in the transition zone. The multivariable model uses compartmental *T_2_* measurements from each voxel in addition to the RSI restriction score to determine the probability that it contains csPCa.

### Voxel-level cancer detection

For voxel-level cancer detection, the 10-fold cross-validation mean AUC of the LRM was 0.98 [95% CI: 0.957, 0.985], versus 0.98 [95% CI: 0.958, 0.986] for maximum RSIrs, indicating that incorporating compartmental *T_2_* did not improve discrimination over RSIrs alone (*p*-value = 0.9).

### Patient-level cancer detection

For cohort 1, the AUC of the LRM was 0.804 [0.648,0.930], versus 0.805 [0.648, 0.931] for RSIrs. The mean AUC for maximum *C_1_* was 0.695 [0.530, 0.851]. The difference in AUCs between RSIrs and the multivariable model was not significantly different (*p*-value = 0.26). Both RSIrs and the LRM performed significantly better than diffusion (maximum *C_1_*) alone (both *p*-values << 0.001).

For cohort 2, the mean LRM AUC for 10,000 bootstrapped samples was 0.724 [0.650, 0.793]. The mean AUC of RSIrs for 10,000 bootstrapped samples was 0.725 [0.652, 0.794]. For maximum *C_1_* the mean AUC was 0.654 [0.573,0.730]. The difference in AUCs between RSIrs and the multivariable model was not significantly different (*p*-value = 0.54). Both RSIrs and the LRM performed significantly better than diffusion (maximum *C_1_*) alone (both *p*-values << 0.001).

**Figure 5:**
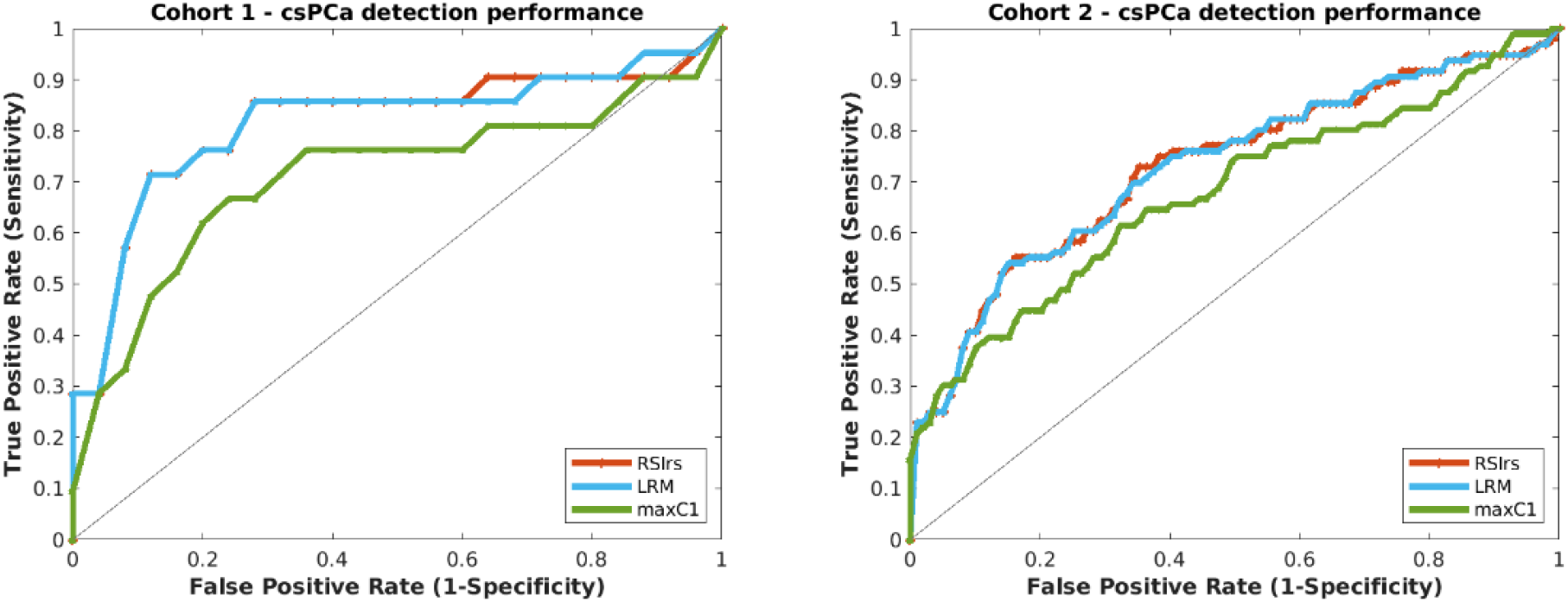
Patient-level diagnostic performance of maximum RSIrs, the LRM and maximum *C_1_* (max *C_1_*) for cohorts 1 and 2. For cohort 1, mean AUC values were 0.695 for maximum *C_1_*, 0.805 for RSIrs and 0.804 for the multivariable model. For cohort 2, mean AUC values were 0.654 for maximum *C_1_*, 0.725 for RSIrs and 0.724 for the multivariable model.

## Discussion

We found that compartmental *T_2_* values were significantly different across RSI diffusion compartments. Moreover, compartmental *T_2_* values differed between csPCa lesions and benign tissue or low-grade PCa in compartments C*_1,_* C*_2,_* and C*_3_*. At the patient level, there were also differences in whole-prostate *T_2_* between patients with no csPCa and those with biopsy-proven csPCa. Quantitative differences in compartmental *T_2_* may provide insight into the microstructural changes associated with PCa. For example, extracellular matrix remodeling may contribute to the increased compartmental *T_2_* observed in *C_2_*^12^. Lower compartmental *T_2_* values in C*_3_* may reflect hyperplasia-induced reductions in luminal space.^13^ While the elevated compartmental *T_2_* observed in *C_1_* of csPCa patients may seem opposed to the known overall hypointense appearance of csPCa on *T_2_*-weighted MRI, it is consistent with an increase in nuclear volume fraction that is typical of cancer cells^24^.

We have previously shown that RSIrs is a useful quantitative DWI biomarker for csPCa at the voxel and patient levels^11,20,22^. Here, we evaluated whether incorporating compartmental *T_2_* values would improve csPCa discrimination. We demonstrated that *T_2_* effects have csPCa discriminatory value, as they showed higher AUC values compared to diffusion alone in both cohorts. However, consideration of compartmental *T_2_* did not significantly improve csPCa-detection performance over maximum RSIrs at the voxel- or patient-level. RSIrs already includes information related to global *T_2_* signal in the prostate (median signal in the whole prostate on the *b*=0 acquisition). It appears that this global *T_2_* assessment may contain sufficient information to normalize RSI diffusion signal and improve csPCa discrimination. This result suggests that compartmental *T_2_* mapping may not be worth the additional scan time and computation required.

Other advanced imaging techniques have also been used to measure *T_2_* in the prostate. LWI utilizes the unique *T_2_* relaxation rates associated with various components of prostate tissue to quantify the fractional volume of glandular lumen, denoted as luminal water fraction (LWF)^25^. This method takes advantage of the observed alterations in the composition of prostatic tissue in the presence of cancer and the Gleason grade of the cancer for PCa diagnosis^26^. Hybrid multidimensional MRI (HM-MRI) exploits the interdependence of *T_2_* and ADC values to measure prostate volume fractions of the lumen, epithelium, and stroma^27^. Studies using LWI and HM-MRI demonstrate lower *T_2_* values in cancer lesions compared to normal prostate tissue with increasing Gleason Grade. The decrease in *T_2_* results from a decrease in luminal volume due to cellular hyperplasia^26,28^. This trend agrees with the decrease observed in this study of *T_2_* in compartment *C_3_* of patients with csPCa. This compartment reflects signal from freely diffusing water in the prostate, which we expect to find predominantly in luminal tissue and to be impacted by a reduction in luminal space. Prior work with HM-MRI showed that the decrease in luminal space is largely the result of epithelial tissue proliferation, indicated by an increase in the measured epithelial volume fraction^27^. Since RSI does not explicitly assign signal contributions to a particular tissue type, this aspect of HM-MRI is harder to align with the present study. However, we can be sure that the changes observed in the *T_2_* of compartment *C_1_* reflect, at least in part, an increase in the overall tissue cellularity^29^. Signal contributions in this compartment are also dependent upon the nuclear volume fraction of cells in the tissue^24^, and the increase in the *T_2_* observed in *C_1_* of csPCa patients suggests an increase in nuclear volume fraction. Whole-mount histopathology data is currently being collected as part of an ongoing study to map changes in compartmental RSI signal to the histological restructuring of prostate tissue due to csPCa.

Future work will also evaluate larger patient cohorts to consider the potential impact of compartmental *T_2_* measurements on the prediction of csPCa grade and tumor extent with the RSI framework. Finally, an ongoing, prospective trial is evaluating the impact of RSIrs and compartmental *T_2_* on csPCa diagnosis in multiple centers and with multiple readers.

### Limitations

The use of separate acquisitions and only two TEs may have limited the accuracy of voxel-wise compartmental *T_2_* measurements. Our study used two TEs for each cohort, where the lowest TE was 76 ms. Other quantitative MRI approaches use lower TEs (less than 30 ms) for *T_2_* mapping^14,26^. The high *b*-values necessary to optimally estimate RSI *C_1_* are generally incompatible with very low TE on clinical scanners, so separate acquisitions would be required to evaluate the combination of optimal RSIrs and *T_2_* estimated with very low TE. Another limitation of this study is that our voxel-level data included only high-confidence csPCa and control categories, leaving little room for improvement over RSIrs for the voxel-level analysis^11^.

## Conclusion

*T_2_* mapping affords insights into characteristics of benign and cancerous prostate tissue, but we did not find compelling evidence that acquisitions with multiple TE is necessary for patient-level csPCa detection with RSI.

## Data Availability

All data produced in the present study are available upon reasonable request to the authors.

